# A Cross-Sectional Survey of the Knowledge, Attitudes & Practices of Antimicrobial Users and Providers in an Area of High-Density Livestock-Human Population in Western Kenya

**DOI:** 10.1101/2021.06.23.21259378

**Authors:** Steven A. Kemp, Gina L. Pinchbeck, Eric M. Fèvre, Nicola J. Williams

**Affiliations:** Institute of Infection, Veterinary and Ecological Sciences, University of Liverpool, UK; Division of Infection & Immunity, University College London, London, UK; Department of Medicine, Cambridge, UK; International Livestock Research Institute, Nairobi, Kenya

**Keywords:** AMR, AMU, veterinary antimicrobials, KAP, antimicrobial stewardship

## Abstract

**Background:** Antimicrobial resistance (AMR) is one of the most important global health crises in recent times and is driven primarily by antimicrobial consumption. In East Africa, there is a paucity of data regarding the knowledge, attitudes and practices related to antimicrobial use (AMU). We investigate the ways in which antimicrobial users in the veterinary sector accessed veterinary antimicrobials, and common behaviours of veterinary antimicrobial users and prescribers associated with AMU and AMR.

**Methods:** In total, 70 farmers, staff at 49 agricultural-veterinary antimicrobial shops (agrovet staff) and 28 veterinary animal healthcare workers or veterinary surgeons (veterinary professionals) were interviewed in Busia county, western Kenya in 2016 using a standard questionnaire as a framework for structured interviews. Data recorded included participant demographics, level of education, access to and sources of veterinary antimicrobials, prescribing patterns and knowledge of AMR and antimicrobial withdrawal periods.

**Results:** The majority of antimicrobials were accessed through informal means, purchased from agroveterinary shops; more than half of staff did not hold nationally mandated qualifications to advise on or sell veterinary antimicrobials. Approximately 40% of veterinary antimicrobials were sold without a prescription and it was noted that both price and customer preference were important factors when selling antimicrobials in almost all agrovet shops. Knowledge of the dangers associated with AMR and AMU were mostly superficial. Treatment failure occurred often, and there was a lack of differentiation between AMR and simply treatment failure.

**Conclusion:** In this study area in East Africa with high-density human and livestock populations, AMU was primarily for maintenance of livestock health. These findings have highlighted several aspects surrounding inappropriate access to antimicrobials, and as such require attention from policy makers concerned with AMR in both livestock and human medicine sectors. Improving prescribing practices and ensuring a minimum level of general education and awareness of prescribers, as well as expanding the role of agrovet staff in antimicrobial stewardship programmes, may help begin to mitigate the maintenance and transmission of AMR, particularly amongst livestock.

## Introduction

Antimicrobials are essential for maintaining animal health in livestock production systems, but inappropriate dispensing and dosing, poor quality of drugs, overuse and self-medication of antimicrobials can select for and exacerbate the emergence, transmission, and persistence of antimicrobial resistance (AMR)^1-3^. In East Africa, there is high demand for animal food products to support the rapidly growing population, and this demand is largely fulfilled by the high proportion (83%) of people engaging in crop and livestock farming^4^. In some parts of the region, such as in the Lake Victoria crescent ecosystem, increased demand has prompted the shift from small holder farming to greater commercialisation and intensification^5^, which often necessitates increased antimicrobial use (AMU) for prophylaxis and treatment of animals in order to maintain animal health^6^. Livestock may act as reservoir of AMR bacteria, with potential for widespread transmission between humans and animals as a result of close contact between the two, or via the food chain. The former is an issue when there are high densities of both humans and livestock^4^, as is the case in both rural and urban Kenya, where this study was conducted^7,8^.

There are significant ramifications of AMR amongst livestock; nine of the 14 classes of antimicrobials considered to be ‘critically important’ for human health are used in both human and livestock health. Three of these (3^rd^-5^th^ generation cephalosporins, fluoroquinolones and polymyxins) are considered to be highest-priority critically important antimicrobials (HPCIAs) for human health^9^. AMU in livestock production is predicted to increase by up to 67% by 2030; as increased AMU may result in significant negative impacts on animal welfare and food security, as well as reducing efficacy of antimicrobials which have crossover for human health^10^. However, it is important to note that owing to the complex epidemiology of AMR, the quantifiable contribution that AMU in livestock has on the emergence, transmission, and maintenance of AMR in humans is still debatable. Studies have shown that similar strains of AMR bacteria are found in both food animals and humans^11^, as well as plasmid-mediated resistance in *E. coli* to polymyxins (mcr-1), originating from food animals^12^. Despite this, other argue that transfer of animal to human resistance genes is negligible and that reduction of AMU in food-producing animals may have a negative effect on food safety and human health^13^. Regardless of the debate, such data to is mostly absent in sub-Saharan Africa.

In many sub-Saharan African countries, including Kenya, there is a paucity of data on the prevalence of both AMR and AMU, as the combined realities of underfunded veterinary healthcare systems, limited regulatory capacities and lack of systematic, national, or regional surveillance systems undermine efforts to promote prudent AMU and control AMR^14,15^. Indeed, Kenya is part of a global effort to improve surveillance capacity in line with its National AMR Action Plan.

Many existing studies examining antimicrobial treatment patterns typically rely on self-reported data, showing that antimicrobials are almost always purchased without prescriptions at ‘agrovets’ (shops which stock agricultural and veterinary antimicrobials as well as other agro-veterinary products)^16-19^. Agrovets are often staffed with pharmaceutical technicians^20^ who have obtained formal training in animal sciences. As such ‘agrovet staff’ may sell antimicrobials, but crucially cannot prescribe them. To comply with local law, agrovet owners may be veterinarians and would thereby be able to legally prescribe antimicrobials. Private veterinary professionals travel to farms at the request of farmers where they provide advice, treat animals, or prescribe veterinary drugs. Veterinary professionals would typically have professional qualifications specifically enabling them to prescribe veterinary antimicrobials and are governed by the Veterinary Surgeons and Veterinary Paraprofessionals Act of the Government of Kenya^21^. Together, antimicrobial sellers and prescribers are responsible for, and play a pivotal role in, highlighting issues that surround AMU and AMR, as well as being the front line of antimicrobial stewardship^22^. Relatively few studies^23^ have examined the knowledge, attitudes, and practices (KAP) of antimicrobial users and prescribers, and such studies are critically required in order to identify risky behaviours and target them for intervention.

In this study, we assessed the way in which antimicrobials were accessed and the general awareness and common behaviours relating to antimicrobial purchase and prescription amongst farmers, agrovet shop staff and veterinary professionals in a small holder livestock production system in western Kenya^24^.

## Methods

### Study Area & Population

A cross-sectional study investigating how farmers, agrovets and veterinary paraprofessionals access and prescribe antimicrobials was conducted in Busia county, western Kenya in 2017. The region was selected for study as it supports the highest human and animal population densities in eastern Africa with approximately 893,681 people^25^, 83% of which engage in livestock production^4^; the region is also broadly representative of other communities spanning the Victoria Lake Basin in Kenya, Uganda, and Tanzania.

Busia county is sub-divided into seven ‘sub-counties’. Within each sub-county, ten farms were randomly selected for interview as a convenience, but also to capture the spatial distribution and diversity of farming practices across the county. Systematic interviewing of agrovet shops and veterinary professionals (**Figure 1**) was conducted with assistance from the sub-county veterinary officer from each sub-county. Interviews were sought with the most senior member of staff in all locatable agrovet shops in the county, except when shops were closed on more than two occasions during repeat visits. A comprehensive list of all known veterinary professionals was collected from sub-county district officers and veterinary professionals were recruited by phone. Veterinary professionals were who agreed to participate were interviewed separately from agrovet shops, at a convenient location to each participant.

**Figure 1.**
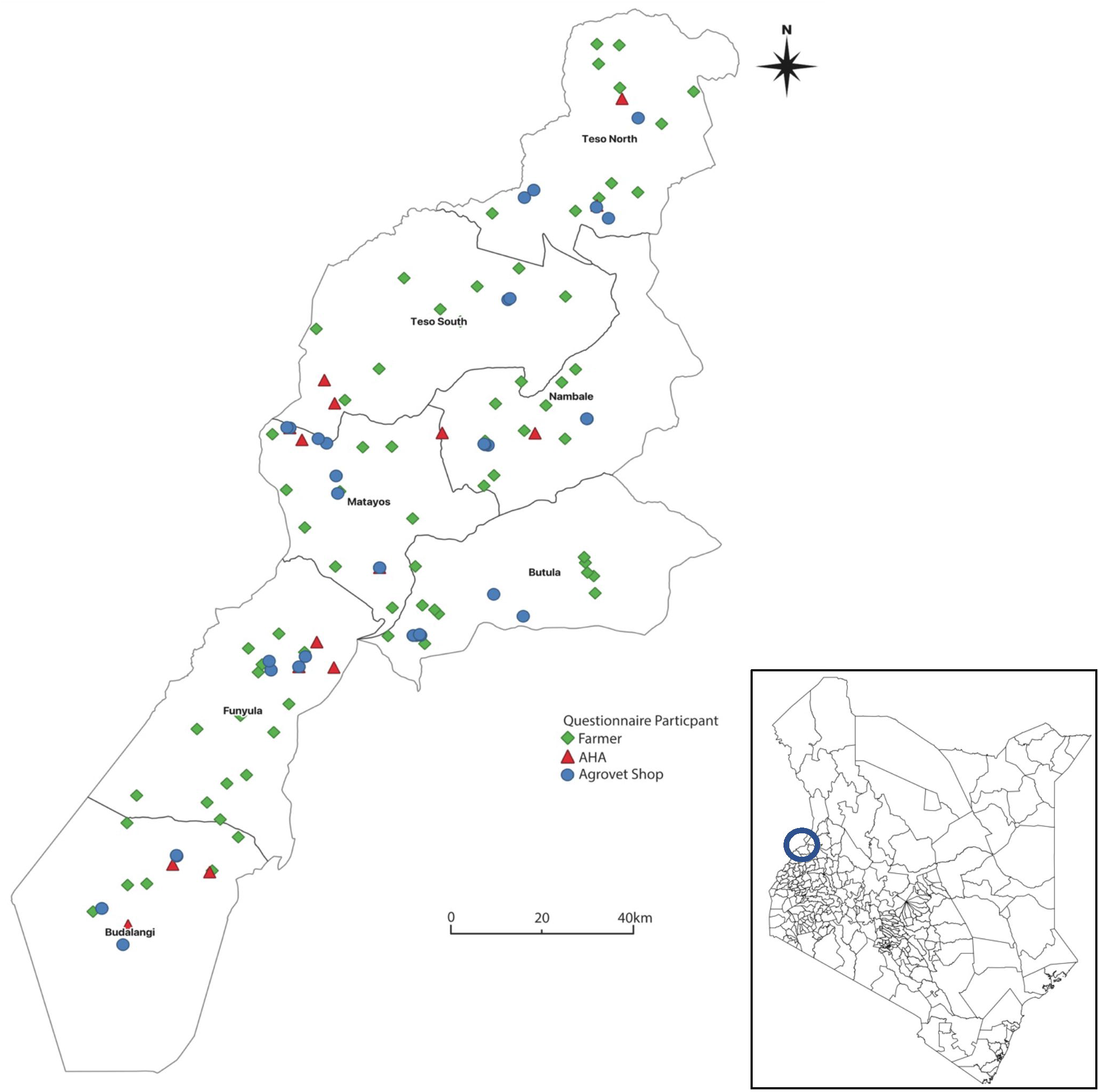
Map of Busia county, western Kenya, indicating the locations of all interviewed farmers, agrovet staff (within their shops) and animal healthcare assistants.

### Questionnaire Design & Piloting

All recruited participants were interviewed orally using a questionnaire as a framework. Questionnaires were designed in Adobe^®^ Acrobat^®^ Pro DC (Adobe, San Jose, United States) and coded electronically using AppSheet^®^ (AppSheet c/o Solvebot Inc., Seattle, Washington). Participants were interviewed in English or Kiswahili by bilingual Kenyan research members. Answers were given verbally by the participant and recorded verbatim as transcribed text into the coded questionnaire on a mobile phone or tablet, by the interviewer. Questions were designed to determine the participant’s education level, access to veterinary antimicrobials, prescribing patterns of antimicrobials, knowledge of antimicrobials, resistance, and withdrawal periods. Questions specifically asked of farmers focused on access to veterinary antimicrobials, basic information on animals kept (date of acquisition, vaccination status), common diseases and understanding of AMR and withdrawal periods. Veterinary professional and agrovet staff questionnaires focused primarily on sales/prescription patterns and responsible use of antimicrobials.

Questionnaires were piloted on field team staff. Minor refinements to question wording were made to better reflect local conditions before conducting a further pilot on a sub-county veterinary officer, before applied in the field. A summary of all questions is presented in (**Supplementary Table 1**).

All questionnaire data can be found at https://github.com/Steven-Kemp/Kenya_KAP.

### Data Analysis

Transcribed answers for each question were imported into Microsoft Excel 2016 (Microsoft Corporation, Redmond, USA). Descriptive analysis including frequencies and percentages for categorical variables (gender, age, education level) were calculated using SPSS Statistics v25.0 (IBM SPSS Statistics for Windows Version 25.0, (New York: IBM Corp). Open-ended questions were analysed on a question-per-question basis using a thematic approach^26^. Briefly, all responses were imported into excel and read twice for familiarisation, data were coded, and then individual themes were generated and checked independently. Finally, themes were reviewed once again, refined, and then presented. Also using SPSS v25.0, the Fisher’s exact test was used to compare specific training undertaken by antimicrobial providers relating to antimicrobial prescription.

Maps were constructed using QGis v3.10 (QGIS Development Team, http://qgis.osgeo.org/). Figures were constructed in Prism v9.1.1.

### Ethical Approval

Ethical approval was obtained from the Institutional Research Ethics Committee of the International Livestock Research Institute (ILRI-IREC2016-03), and the University of Liverpool Veterinary Science Research Ethics Committee (VREC387). All participants gave informed, written consent before participation in the study.

## Results

### Participant Demographics & Education

A total of 70 farmers, 49 staff at agrovet shops, 27 AHAs and 1 veterinary surgeon were recruited (**Table 1**). As recognised professionals, the veterinary surgeon and AHAs were considered together in our analysis and are referred to as ‘veterinary professionals’ throughout. The predominant age bracket for all groups surveyed was 25-44. The majority of agrovet staff were either agrovet assistants (79.6%) or shop owners (18.4%). Only 44.9% of agrovet staff had obtained college or university education, compared to 89.2% of veterinary professionals. For farmers, the majority (47.1%) had completed at least secondary school education. Significantly more veterinary professionals had received specific training in livestock health and disease (*P* = 0.01) than agrovets. Only 42.9% of agrovets and 82.1% of veterinary professionals had received specific training to dispense veterinary antimicrobials. A large proportion of agrovets cited informal training (44.9%) as their primary source of knowledge, compared to 92.9% of veterinary professionals who obtained a professional qualification awarded by a college or university. However, 7.1% (n=2) of veterinary professionals interviewed stated that they did not have university education, therefore could not be called veterinary professionals.

**Table 1.** Participant Demographics and Education

| Characteristics |  | Agrovet Staff<br>(n=49) |  | Veterinary<br>professionals (n=28) |  | Farmers<br>(n=70) |  |
| --- | --- | --- | --- | --- | --- | --- | --- |
|  |  | n | % | n | % | n | % |
| **Gender** | Male | 25 | 51.0 | 27 | 96.4 | 48 | 68.6 |
|  | Female | 24 | 49.0 | 1 | 3.6 | 22 | 31.4 |
| **Age Group** | 18-24 | 8 | 16.3 | - | - | 2 | 2.9 |
|  | 25-44 | 35 | 71.4 | 20 | 71.4 | 40 | 57.1 |
|  | 45-64 | 5 | 10.2 | 6 | 21.4 | 17 | 24.3 |
|  | 65+ | 1 | 2.0 | 2 | 7.1 | 11 | 15.7 |
| **Job Position** | Animal Healthcare Worker | 1 | 2.0 | 14 | 50.0 | - | - |
|  | Artificial Insemination Technician | - | - | 1 | 3.6 | - | - |
|  | Sub-country Veterinary Officer | - | - | 3 | 10.7 | - | - |
|  | Agrovet Assistant | 39 | 79.6 | 1 | 3.6 | - | - |
|  | Laboratory Staff / Vet Technician | 1 | 2.0 | 3 | 10.7 | - | - |
|  | Livestock Officer | - | - | 5 | 17.6 | - | - |
|  | Veterinarian | - | - | 1 | 3.6 | - | - |
|  | Manager | 1 | 2.0 | - | - | - | - |
|  | Owner | 9 | 18.4 | - | - | - | - |
| **Length of time at job** | <1 Year | 14 | 28.6 | 1 | 3.6 | - | - |
|  | 1-2 Years | 4 | 8.2 | - | - | - | - |
|  | >3 Years | 31 | 63.3 | 27 | 96.4 | - | - |
| **Highest Education Level** | No Formal Education | - | - | - | - | 4 | 5.7 |
|  | Primary Education | - | - | - | - | 24 | 34.3 |
|  | Secondary Education | 27 | 55.1 | 3 | 10.7 | 33 | 47.1 |
|  | College (certificate/diploma) | 20 | 40.8 | 23 | 82.1 | 7 | 10 |
|  | University | 2 | 4.1 | 2 | 7.1 | 2 | 2.9 |
| **Nature of Training** | Professional Qualification | 8 | 16.3 | 26 | 92.9 | - | - |
|  | Pharmaceutical company | 15 | 30.6 | - | - | - | - |
|  | None/Informal Training | 22 | 44.9 | 2 | 7.1 | - | - |
|  | Cannot Remember | 3 | 6.1 | - | - | - | - |

### Access to antimicrobials and common sales patterns

All veterinary antimicrobials were purchased directly from agrovet shops, where both farmers and veterinary professionals can purchase antimicrobials from. Antimicrobials and vaccines were distributed to local agrovets shops by two larger wholesale agrovet shops (one within Busia county, one in neighbouring Bungoma county) who obtained antimicrobials directly from manufacturers and through their supply chains.

Farmers reported no restrictions (in amount or class) when purchasing antimicrobials from agrovet shops, even without a valid prescription. More than half (57.1%) of veterinary professionals stated that they provided a prescription for farmers to obtain antimicrobials, with the remainder treating animals with their own stock and billing farmers separately for these. This agreed with responses from agrovet staff who reported that they (60%) often dispensed antimicrobials against a prescription. Direct observations when visiting such premises confirmed that agrovet shops did sell antimicrobials with no prescription, as well as dispensing single syringes of formula antimicrobials or partial-treatments to farmers, even though this is a contravened practice in Kenyan Law^27^.

Participants were asked to indicate the most commonly sold or prescribed antimicrobials (agrovet staff and veterinary professionals) or most commonly purchased (farmers), and a total of 26 different antimicrobials were reported by all groups. Oxytetracycline and penicillin-streptomycin were the two most commonly sold or prescribed antimicrobials by agrovet staff and veterinary professionals (**Table 2**), followed by sulfonamides. The majority of farmers opted to purchase oxytetracycline as their primary drug of choice (78.6%) from agrovet shops. There was no reported use or sale/prescription of 3^rd^+ generation cephalosporins or fluoroquinolones. There was only a single occasion whereby a farmer purchased polymyxins (colistin), but these drugs are available at agrovets when requested.There were large inconsistencies in the reported use of antimicrobials. Antimicrobials were predominantly reported as being used therapeutically (i.e. not for growth promotion or prophylaxis) by farmers (85.7%) and veterinary professionals (100%), and sold for therapeutic purposes by agrovets (98.0%). However, prophylactic use of antimicrobials was subsequently indicated by 37.1% of farmers and 28.6% of veterinary professionals and sold as such by 38.8% of agrovets in a later question in the questionnaire. Use of antimicrobials as growth promoters was reported by 37.1% farmers, but not sold as such by agrovet shops or prescribed by veterinary professionals.

**Table 2.** List of the most commonly used/purchased/prescribed antimicrobials according to farmers, agrovets and veterinary professionals, to treat livestock. Up to 5 ‘most common’ antimicrobials were volunteered; therefore, each antimicrobial was counted once each time it featured in the respondents’ answer.

| Antimicrobial | Veterinary professionals (n=28) |  | Agrovet Staff (n=49) |  | Farmers (n=70) |  |
| --- | --- | --- | --- | --- | --- | --- |
|  | n | % | n | % | n | % |
| Oxytetracycline | 26 | 92.9 | 46 | 93.9 | 55 | 78.6 |
| Penicillin-streptomycin | 27 | 96.4 | 39 | 79.6 | 33 | 47.1 |
| Sulfachloropyrazine | 9 | 32.1 | 27 | 55.1 | - | - |
| Sulfadimidine | 9 | 32.1 | 13 | 26.5 | 2 | 2.9 |
| Trimethoprim & Sulfadiazine | 9 | 32.1 | 8 | 16.3 | 4 | 5.7 |
| Tylosin & Doxycycline | - | - | 18 | 36.7 | 2 | 2.9 |
| Sulfamethoxazole | 3 | 10.7 | 8 | 16.3 | - | - |
| Gentamicin | 6 | 21.4 | - | - | 1 | 1.4 |
| Tylosin | 4 | 14.3 | - | - | - | - |
| Tetracycline | 1 | 3.6 | 3 | 6.1 | - | - |
| Fosfomycin & Tylosin | - | - | 4 | 8.2 | - | - |
| Sulfamethoxazole & Trimethoprim | - | - | 4 | 8.2 | - | - |
| Erythromycin | 2 | 7.1 | - | - | 1 | 1.4 |
| Gentamicin & Doxycycline | - | - | 3 | 6.1 | - | - |
| Neomycin | - | - | 3 | 6.1 | - | - |
| Cefalexin | 1 | 3.6 | - | - | 1 | 1.4 |
| Metronidazole | 1 | 3.6 | - | - | 1 | 1.4 |
| Ampicillin | 1 | 3.6 | - | - | - | - |
| Streptomycin | 1 | 3.6 | - | - | - | - |
| Amoxicillin | - | - | 1 | 2.0 | - | - |
| Dexamethasone** | - | - | 1 | 2.0 | - | - |
| Erythromycin & Oxytetracycline | - | - | 1 | 2.0 | - | - |
| Colistin* | - | - | - | - | 1 | 1.4 |

\*Highest Priority Critically Important Antimicrobials \*\*not an antimicrobial but described by the respondent as one.

The most common diseases that antimicrobials were cited as being purchased to treat were East Coast fever (theileriosis), anaplasmosis, trypanosomiasis, diarrhoea, and general respiratory diseases.

### Advice and considerations given at point of sale regarding AMU, AMR, and withdrawal periods

Most farmers reported first seeking the advice of a veterinary professional before purchasing antimicrobials (78.6%). More than half of farmers (54.3%) never requested specific antimicrobials without first discussing with either agrovet staff or veterinary professional. A small minority of farmers (12.9%) purchased antimicrobials without obtaining any advice from an agrovet or a prescription from a veterinary professional. Such farmers stated they did so “using [their] own knowledge” or “already had a prescription from a veterinary officer from a previous consultation”. A small proportion of farmers also reported using antimicrobials previously prescribed or purchased, “[having antimicrobials leftover] from previous use”.

In agrovet shops the primary consideration when selling antimicrobials was customer preference (65.3%). Veterinary professionals’ primary consideration was antimicrobial effectiveness (57.9%) and then cost (39.3%). Farmers were primarily concerned with antimicrobial cost (44.3%), followed by effectiveness (40.0%). As cost was a common consideration, the sale price of various antimicrobials was collected (**Figure 2**). The average price of oxytetracyclines were cheaper than penicillin/streptomycin; this is consistent with the finding that oxytetracyclines were the most commonly sold antimicrobial in agrovets shops. A small minority of farmers also considered antimicrobial availability and the distance they needed to travel to purchase specific types of antimicrobials as their primary point of consideration (5.7%). Specific agrovet shops were chosen by farmers for several reasons including the “close distance to [their] farms”, ability to “get drugs on credit” and for “wide selection” and “good stock availability”.

**Figure 2.**
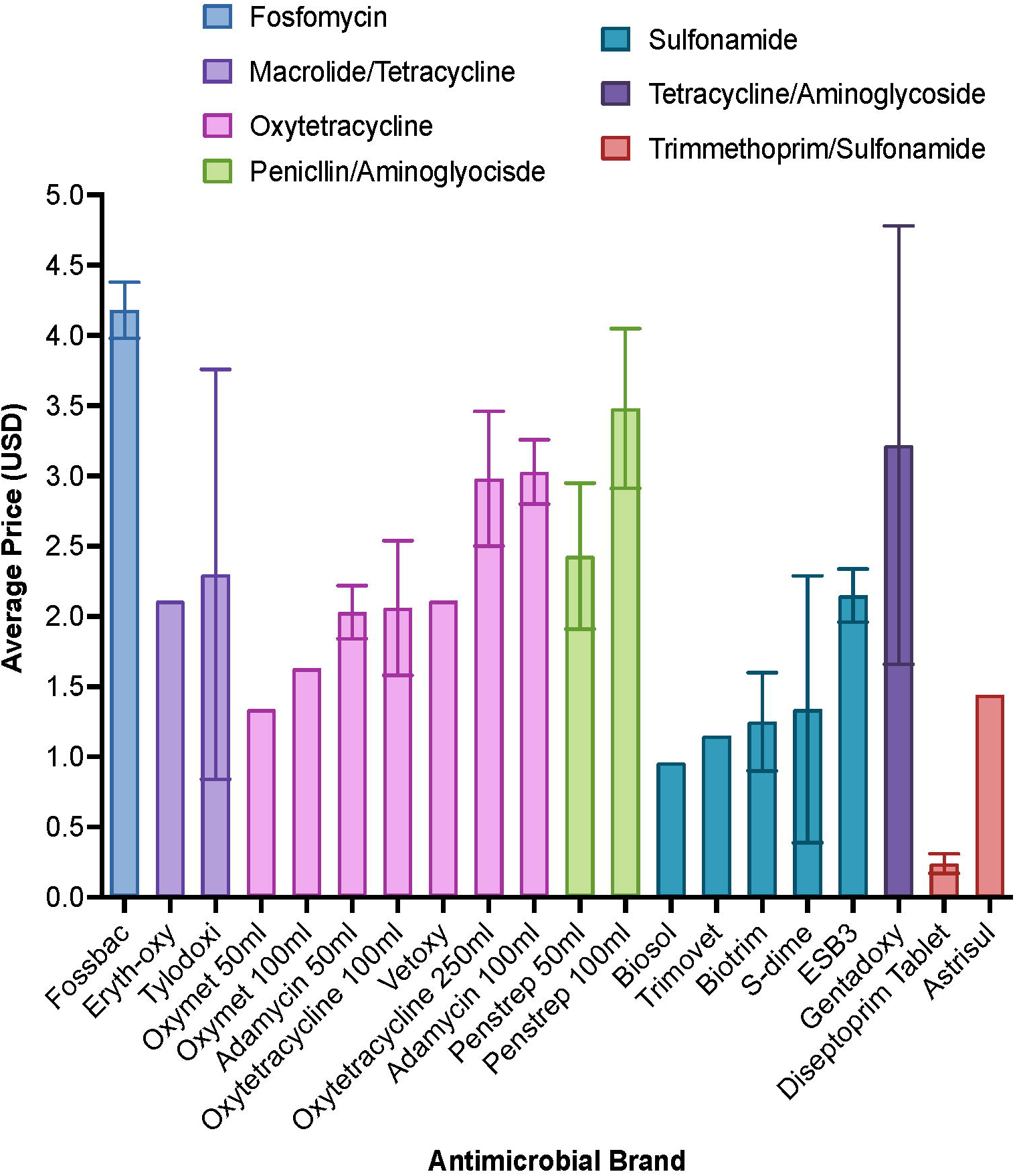
Average sale cost of antimicrobials from 49 agrovet shops across Busia county. Error bars represent standard deviation where more than one agrovet shop reported pricing data.

The most commonly offered information regarding antimicrobials at point of sale or prescription differed significantly between antimicrobial sellers and antimicrobial providers; 61.2% of agrovet staff gave directions for use of antimicrobials, compared to only 25.0% of veterinary professionals, where they were provided to the farmer to use themselves. Similarly, significantly more veterinary professionals chose to give no information at all (50.0%) compared to 18.4% of agrovets (**Figure 3**). The other two most common cited pieces of information provided to farmers were withdrawal periods and dosage instructions, though in all cases, these were reported to be read from the packaging.

**Figure 3.**
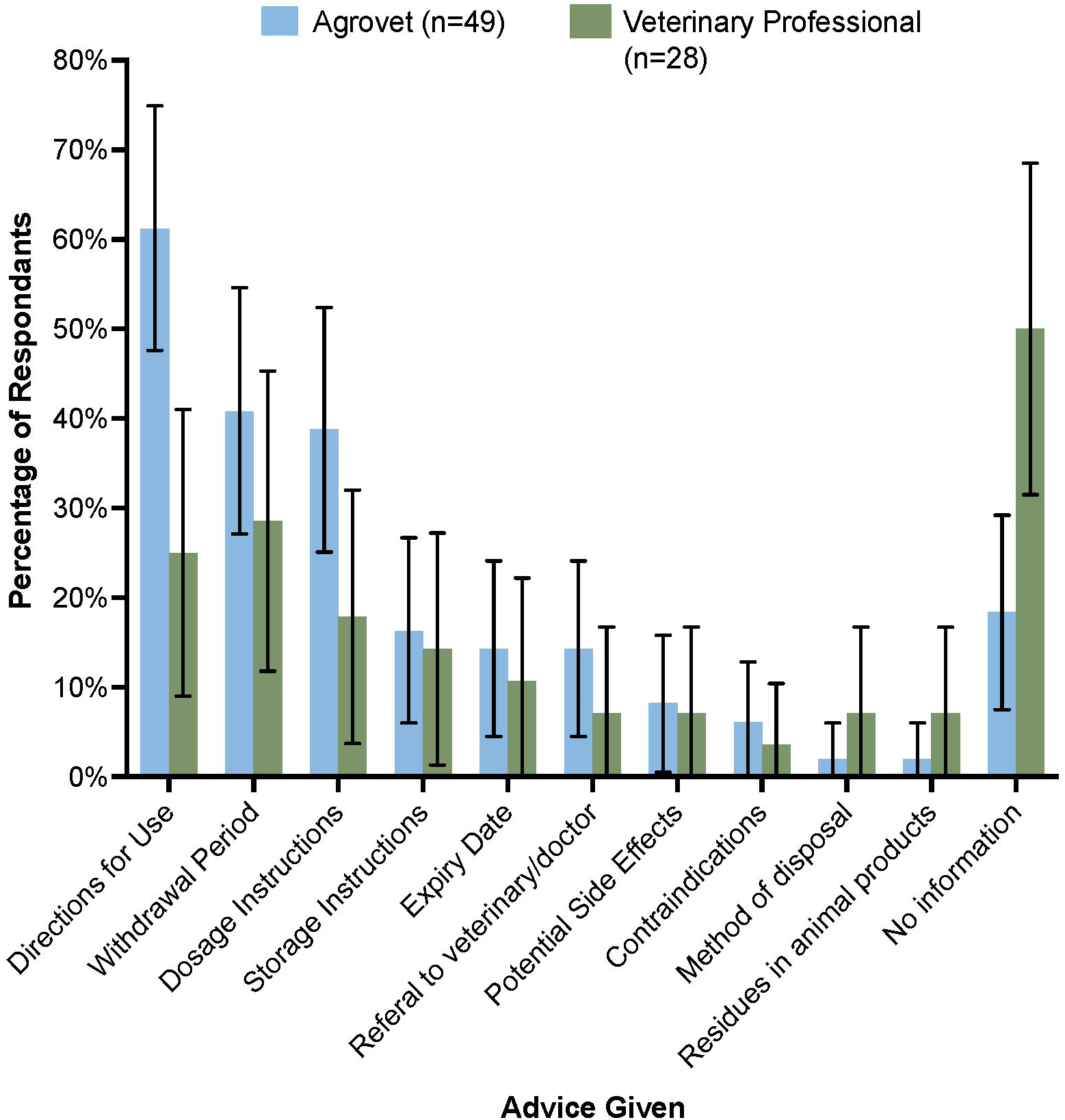
Information given to farmers regarding AMU, AMR and withdrawal periods at point-of-sale (agrovet shop) or when receiving a prescription (veterinary professional). Error bars represent 95% confidence intervals.

### Understanding of AMR

Participants rarely recognised the terms “antimicrobial resistance” or “antibiotic resistance”. Once given a definition, many suggested that they had heard of it, but did not recognise the specific terminology. A large proportion of agrovets (69.4%), veterinary professionals (39.3%) and farmers (47.0%) did not know the causes of AMR. Of those who had some knowledge of causes, the most common response was underdosing (significantly more veterinary professionals than agrovets) and prolonged use (**Figure 4**). Some farmers additionally reported “bacteria mutation” (2.9%), “misdiagnosis by an agrovet/veterinary professional” (15.9%) and using “counterfeit antimicrobials” (1.4%) as causes of AMR. Participants who were unsure about the cause of AMR instead guessed: “when you treat an animal and it doesn’t respond”, “when the animal is tired, the antibiotic will not work” and “cheap drugs no longer work, but the more expensive ones do”. Of those respondents who were familiar with AMR, they suggest that there may be resistance to oxytetracyclines, penicillin-streptomycin and sulfonamides were posited, though no formal resistance testing was routinely undertaken.

**Figure 4.**
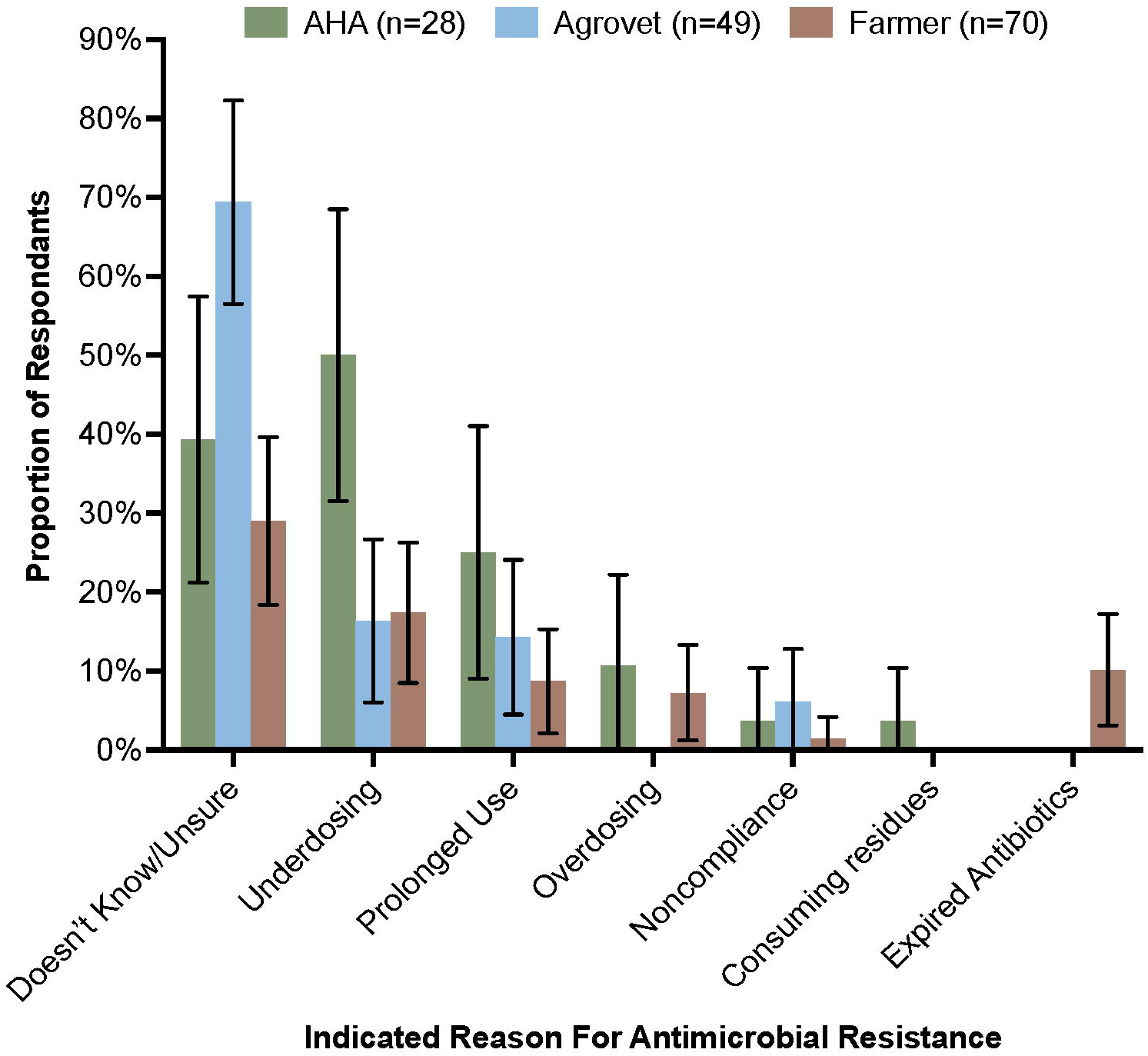
Most common responses given by participants indicating what they thought were the main causes of antimicrobial resistance. Error bars are 95% CI.

Knowledge of withdrawal periods was mostly superficial amongst farmers. Contrary to EU regulations, withdrawal periods are usually specific to the route of administration e.g. antimicrobials administered to cattle may have a nil milk withdrawal due to penetration into the udder, but would have a meat withdrawal period – this is not often defined on antimicrobial packaging (**Supplementary Figure 1a-d**). However, with respect to withdrawal periods or definitions, 12.9% having “no understanding” (never heard of withdrawal period before), 34.3% had “minor understanding” (had heard of it but quoted incorrect withdrawal periods for animal food products), and 27.1% had “good understanding” (good knowledge and accurate recall of withdrawal periods of each antimicrobial they regularly treated animals with). The remainder (18.6%) stated they sometimes referred to antimicrobial packaging for withdrawal period times. The majority of farmers stated that they did not sell or consume animals or animal products during withdrawal periods (75.7%), though some reported that they purposefully chose to ignore withdrawal period recommendations (17.1%). Commonly farmers fed antimicrobial residue-containing milk to their dogs (14.3%) or allowed calves to suckle during treatment (44.3%). One farmer stated that they regularly gave contaminated milk to their animals, despite understanding the danger of consuming residues: “[I] give to the calves and the dog. [I] understand that resistance may develop in these animals, but [I] choose to ignore it to avoid waste”.

### Management of Drug Failure

Only one instance of a highest-priority critically important antibiotics (HPCIA) was reportedly sold or purchased during the study - colistin. No agrovet staff and only a veterinary professional (3.6%) had heard the term “HPCIA” before. The majority of veterinary professionals and agrovets were unaware of any specific guidelines for antimicrobial prescription or sale, which also extended to sale and use of HPCIAs. Some veterinary professionals cited guidelines from the Kenya Veterinary Board (21.4%) or instructions from the County Veterinary Officer (10.7%) regarding sales or use of antimicrobials. Agrovet staff cited pharmaceutical guidelines (6.1%) or Kenya Veterinary Board guidelines (14.3%).

In terms of defined AMR, there were no confirmed instances due to no diagnostics. However, few instances of clinical failure were reported by agrovet staff (by proxy of farmers returning to purchase an alternative antimicrobial from them). Where clinical failure was reported, reported failures were to oxytetracyclines (10.2%), penicillin-streptomycin (4.1%) and sulfonamides (8.1%). The majority of agrovets indicated that they “[did] not know” or there was “no reported” resistance to antimicrobials (61.2%). Some stated that there had been cases of suspected clinical failure attributed to AMR, but they did not know to which antimicrobial (16.3%) and this was not verified in a laboratory setting. Veterinary professionals suggested that some clinical failures may be attributed to AMR, and that such failures occurred in oxytetracycline (41.4%) and penicillin-streptomycin (27.6%), but not to sulfonamides. Farmers suggested that they had encountered treatment failure in less than half of cases (41.3%). Of those who reported failure, oxytetracycline was the most common (20.6%), followed by penicillin-streptomycin (7.9%) and sulfonamides (1.6%). A small subgroup of farmers suggested that there had been failure, but were unsure to which antimicrobial (11.1%).

Where there was treatment failure, approximately half of veterinary professionals reportedly collected a venous blood smear (53.6%) or sent blood for a bacterial culture (7.1%), or PCR (3.6%). The remainder prescribed an alternative antimicrobial without conducting diagnostics. A quarter of agrovet staff involved a more experienced agrovet staff member or veterinary professional, or the owner of an agrovet shop (28.6%) where they received a report of treatment failure. More than a quarter (26.5%) would suggest an alternative antimicrobial without gaining more information regarding the animal and 22.4% had not encountered treatment failure before. The remainder of agrovet staff would first try to obtain more information i.e. ask about more clinical signs, and then recommend an alternative antimicrobial.

Many antimicrobials prescribers/sellers (64.3% of veterinary professionals and 71.4% of agrovet shops) kept some form of records regarding antimicrobial sale or prescription or incidence of treatment failure. There was good concordance between antimicrobials volunteered as regular purchases or prescriptions and those records that we read. Half of farmers (50.0%) also had some records of antimicrobials they administered to their animals though these were often nonspecific i.e. did not often contain specific antimicrobial names or dosages. When questioned, farmers were often unsure which antimicrobials were used as a veterinary professional had provided and administered the treatment, and not recorded it for them (corroborating the previous point that veterinary professionals do not provide detailed information regarding antimicrobials to farmers).

## Discussion

Our study has shown that all interviewed farmers and veterinary professionals in Busia county accessed veterinary antimicrobials through agrovet shops and that there were, in practice, no restrictions on class or quantity that could be purchased. The most commonly purchased veterinary antimicrobials were tetracyclines, sulfonamides and penicillins. This study found no reported use of fluoroquinolones or 3^rd^+ generation cephalosporins, and only one reported use of colistin; given that these antimicrobials are critically important for human health^9,28^ this was a positive finding. However, few veterinary professionals or agrovet staff recognised examples of HPCIAs despite being presented with a list of those antimicrobials - this is likely due to lack of awareness and available information. Interestingly, these drugs are widely available and found to be drugs of choice amongst some farmers in more urbanised areas of Kenya^7^, despite their relatively higher cost.

Our study highlighted a number of poor antimicrobial-related sale practices in agrovet shops, notably the dispensing of antimicrobials without a prescription and the inclusion of customer preference as a primary consideration when selling antimicrobials. Approximately 40% of agrovet staff stated that they dispensed antimicrobials without a valid prescription, though direct observations made during the study suggested that all shops sold antimicrobials without a prescription at least occasionally; this is consistent with similar studies conducted in Nairobi^23^ and Tanzania^29^. Indeed, observations made during the study also suggest that there is a lack of formal written prescriptions, and that most prescriptions are simply verbal instructions from a veterinary or veterinary paraprofessional. However, this is potentially for convenience, where travel to an agrovet shop or a veterinary professional cannot travel, or a farmer cannot afford to pay for an in-person visit, to a farm. Furthermore, there were significant inconsistencies in the reported use of antimicrobials. Despite not being prescribed or sold as such, antimicrobials we suggest that antimicrobials were used for prophylaxis and/or growth promotion based on participant responses.

In our study, cost of antimicrobials to farmers was a major consideration – we noted that oxytetracyclines were on average cheaper than penicillin-streptomycin (relative to number of doses per container), but more expensive than sulfonamides (**Figure 2**). To save on costs, farmers, who in this study represent a low income group^30^ sometimes opted to bypass veterinary professionals when treating their animals. Numerous farmers stated that they had reused prescriptions from a previous encounter with a veterinary professional or agrovet staff, or they opted to use leftover antimicrobials from previous treatment because they had previously worked. To prevent such irrational drug use by farmers, 75% of veterinary professionals purposely did not provide any direction for antimicrobial use to farmers so that full responsibility for treating animals remained with them (**Figure 3**). Farmer administration of drugs would be difficult to control; this would require interventions that limit access to antimicrobials^31^ but also would require regulation of pricing structures for access to veterinary care, which might be challenging in a liberalised veterinary market^32^.

One of the major drivers of AMU is commercial gain. Livestock production is an important industry in developing countries, driven by market demand and financial incentives. As such, farmers need to keep their animals healthy and resort to this by using antimicrobials. In Kenya, antimicrobials are viewed as high-margin products that are typically administered or sold by a recognised professional. Agrovet staff are routinely approached by large pharmaceutical companies to train staff (**Table 1**) regarding specific antimicrobials they are selling and encourage them to purchase stock for their shops. As staff have made an investment, they would therefore preferentially sell these antimicrobials, even in instances where a cheaper antimicrobial may be more appropriate. It is a lucrative business, as is testament to the large number of agrovet shops and informal veterinary antimicrobial sellers found within this KAP study. Separately, veterinary professionals are paid a salary and would make additional money through extension services, such as selling antimicrobials directly to a farmer, and then charging them for administering those antimicrobials (and taking responsibility for treatment and follow-up care of those animals). Where farmers may be unable to afford such services, they would resort to noting what the veterinary professional has done, and attempt to replicate this later, by purchasing antimicrobials without a prescription.

Few studies have focussed on antimicrobial prescribers and sellers and their knowledge of AMR in LMICs^33^. In this study there was mostly superficial knowledge of AMR and the dangers of AMU amongst farmers, agrovet staff and some veterinary professionals (**Figure 4**). This may have been due to specific terminology, as other studies have highlighted that there is minimal familiarity with terms such as AMR and antibiotic resistance^34,35^. After an accurate definition was provided, some interview participants were able to correctly give examples of factors which they thought may contribute to the emergence of AMR. Withdrawal periods were also generally not well understood or abided by. A study conducted in neighbouring Tanzania found that depending on the region, people were variably likely to observe withdrawal periods^36^, highlighting different attitudes to AMR amongst people engaged in different types of agriculture. If there is insufficient knowledge of the contribution of antimicrobial residues, this may indicate why. Some farmers in our study suggested that withdrawal periods only applied to milk or eggs and were unaware that residues may also occur in meat. There is clear scope, in line with Kenya’s AMR Action Plan, to improve knowledge of the livestock keepers and address the poor understanding of rational drug use amongst farmers and antimicrobial sellers; innovative approaches such as information design (which delivers relevant information in an accessible way to the end user)^37^ could play a role in communicating information regarding AMR in appropriate and simple ways.

An important issue identified in this study was ambiguity surrounding AMR. As there is a routine lack of diagnostics undertaken, cases of treatment failure may be attributed to use of incorrect antimicrobials or incorrect dosing, rather than development of AMR. Veterinary professionals typically relied on their clinical experience for disease identification, and agrovet staff relied on farmer description of animal disease, or more experienced agrovet staff to advise on an appropriate treatment for those reported signs. Several diagnostic laboratories exist in western Kenya, though the cost involved in collecting samples, shipping them to a laboratory and the testing itself is a barrier to most farmers, who cannot afford such services. As such, there is over-reliance on empirical, broad-spectrum antimicrobials.

Because AMR surveillance has not been systematically conducted in Kenya, there is incomplete knowledge of the prevalence of AMR and AMU. Whilst other studies have shown a high prevalence of AMR amongst humans and livestock in LMICs^38,39^, data is lacking in Kenya. Absence of documentation regarding veterinary antimicrobial therapies, systematic reporting of treatment failures and AMR surveillance, precludes gaining an accurate representation of issues surrounding AMR in the current circumstances.

There are complex factors at play surrounding antimicrobial prescription, including high public demand for access to antimicrobials. We suggest that several behavioural interventions in tandem with legislative or policy reforms implemented to agrovet, and veterinary professional staff may reduce inappropriate prescription. We suggest three major intervention: 1) Detailed guidance on alternative, non-antimicrobial therapies could be delivered to agrovet shops from local government. In instances where a diagnosis is made by a veterinary professional, consultation of documentation may suggest that an antimicrobial is not generally indicated for that diagnosis and several alternatives may be suggested. 2) Specific justification for prescription of antimicrobials. Where an agrovet or veterinary professional prescribes an antimicrobial, they must explicitly justify why this was necessary and why an alternative therapy could not be used. Previous studies have found that staff accountability significantly improves decision making accuracy^40^. 3) Ranking of veterinary professionals and agrovets. Each sub-county in Busia tracks the number of agrovet shops and registered veterinary professionals; these staff could be ranked depending on the number of inappropriate prescriptions that have been made and sent an email or text message informing them on their prescribing rates, compared to others. Peer comparison is a strong driver of performance and may help to keep inappropriate prescription low, as has been studied in clinical settings elsewhere^41^. Concurrently implementing these interventions may significantly reduce the number of antimicrobials prescribed, whilst also maintaining a high standard of care expected from farmers treating their animals. Finally, to reiterate relevance of antimicrobial stewardship training, studies have shown that in major national referral and teaching hospitals in Kenya, fewer than 15% of clinicians had received substantial lectures on antimicrobial stewardship and AMR during their training^42^. Reform of veterinary and medical certificates, diplomas and undergraduate training as well as continuing professional development should be made to better equip veterinary professionals to deal with AMU, AMR and stewardship. Such interventions could be implanted with ease via the rollout of the new National Action plan on prevention and containment of antimicrobial resistance, being implemented by the Fleming Fund (https://www.flemingfund.org/wp-content/uploads/0cff5e08e6a64fcf93731d725b04792e.pdf)

This study determined that community-owned agrovet shops are the primary level of veterinary care in an area of smallholder crop-livestock farming. Previous studies have shown positive correlations between AMU and the level of AMR in animal populations^43,44^, and therefore, use of antimicrobials in this smallholder farming production may constitute a major contributing factor to the development of AMR. To remedy this, antimicrobial stewardship must be foremost for prescribers and sellers. As well as improving knowledge in the retail and farming sectors, efforts should be made to standardise record-keeping into a computerised system managed in collaboration with local government, to allow for accurate tracking of prescribed and sold antimicrobials and minimise over- and non-prudent use of antimicrobials, whilst factoring in perceived interventions.

## Conclusions

The findings presented in this study suggest that there was low awareness of both AMU and AMR amongst both antimicrobial users and prescribers, which can have significant public health implications. High rates of AMU (and subsequently AMR) will eventually lead to a situations where there is significantly reduced antimicrobial efficacy in both veterinary and human medicine. In particular, inappropriate prescribing practices by agrovet shops highlights the need to encourage diverse forms of targeted education and behavioural interventions, focussed on prudent antimicrobial prescription and use, in combination with the deployment of national level AMR surveillance in both the livestock and human populations utilising an inter-sectoral collaborative approach to restrict the inappropriate use of antimicrobials.

## Data Availability

Original data gathered is published in this manuscript or in supplementary text. Additional data may be requested from the corresponding author and will be disclosed at reasonable request,

## Transparency Declaration

None to declare.

## Data availability statement

Participant response data (minus potentially identifiable data such as location and region) collected as part of this study has been deposited on GitHub (https://github.com/Steven-Kemp/Kenya_KAP).

## Acknowledgements

We would like to thank Jane Poole (International Livestock Research Institute, Nairobi) for her assistance with the study design, the field team at ILRI and Dr Salome Bukache (University of Nairobi) for helping to pilot the questionnaire. We also thank Maseno Cleophas and Maurice Omondi for travelling with us assistance with executing the questionnaire. Thankk you to individual Busia sub-county veterinary officers for accompanying us on field visits. We would also like to thank all questionnaire participants who gave their time for this study.

## References

1 Padget, M., Guillemot, D. & Delarocque-Astagneau, E. Measuring antibiotic consumption in low-income countries: a systematic review and integrative approach. Int J Antimicrob Agents 48, 27–32, doi:10.1016/j.ijantimicag.2016.04.024 (2016).

2 Bebell, L. M. & Muiru, A. N. J. G. h. Antibiotic use and emerging resistance: how can resource-limited countries turn the tide? 9, 347–358 (2014).

3 Jasovsky, D., Littmann, J., Zorzet, A. & Cars, O. Antimicrobial resistance-a threat to the world’s sustainable development. Ups J Med Sci 121, 159–164, doi:10.1080/03009734.2016.1195900 (2016).

4 Statistics, K. N. B. o. (Inst. for Resource Development/Westinghouse, 2015).

5 Chen, M. et al. Diversification and intensification of agricultural adaptation from global to local scales. PloS one 13, e0196392, doi:10.1371/journal.pone.0196392 (2018).

6 Van Boeckel, T. P. et al. Reducing antimicrobial use in food animals. Science 357, 1350–1352, doi:10.1126/science.aao1495 (2017).

7 Muloi, D. et al. Epidemiology of antimicrobial-resistant Escherichia coli carriage in sympatric humans and livestock in a rapidly urbanizing city. Int J Antimicrob Agents 54, 531–537, doi:10.1016/j.ijantimicag.2019.08.014 (2019).

8 Falzon, L. C. et al. One Health in Action: Operational Aspects of an Integrated Surveillance System for Zoonoses in Western Kenya. Front Vet Sci 6, 252, doi:10.3389/fvets.2019.00252 (2019).

9 WHO. WHO list of critically important antimicrobials for human medicine (WHO CIA list). (World Health Organization, 2019).

10 Van Boeckel, T. P. et al. Global trends in antimicrobial resistance in animals in low- and middleincome countries. Science 365, eaaw1944, doi:10.1126/science.aaw1944 (2019).

11 Pirolo, M. et al. Unidirectional animal-to-human transmission of methicillin-resistant Staphylococcus aureus ST398 in pig farming; evidence from a surveillance study in southern Italy. Antimicrobial resistance and infection control 8, 187, doi:10.1186/s13756-019-0650-z (2019).

12 Bich, V. T. N. et al. An exploration of the gut and environmental resistome in a community in northern Vietnam in relation to antibiotic use. Antimicrobial resistance and infection control 8, 1–10, doi:ARTN 194 10.1186/s13756-019-0645-9 (2019).

13 Mather, A. E. et al. Distinguishable epidemics of multidrug-resistant Salmonella Typhimurium DT104 in different hosts. Science 341, 1514–1517, doi:10.1126/science.1240578 (2013).

14 Cox, J. A. et al. Antibiotic stewardship in low- and middle-income countries: the same but different? Clinical microbiology and infection : the official publication of the European Society of Clinical Microbiology and Infectious Diseases 23, 812–818, doi:10.1016/j.cmi.2017.07.010 (2017).

15 Founou, L. L., Founou, R. C. & Essack, S. Y. Antibiotic Resistance in the Food Chain: A Developing Country-Perspective. Frontiers in microbiology 7, 1881, doi:10.3389/fmicb.2016.01881 (2016).

16 Johnson, S., Bugyei, K., Nortey, P. & Tasiame, W. Antimicrobial drug usage and poultry production: case study in Ghana. Animal Production Science 59, 177–182, doi:10.1071/An16832 (2019).

17 Higham, L. E., Ongeri, W., Asena, K. & Thrusfield, M. V. Characterising and comparing animal-health services in the Rift Valley, Kenya: an exploratory analysis (part I). Tropical animal health and production 48, 1621–1632, doi:10.1007/s11250-016-1136-0 (2016).

18 Mboya, E. A., Sanga, L. A. & Ngocho, J. S. Irrational use of antibiotics in the Moshi Municipality Northern Tanzania: a cross sectional study. The Pan African medical journal 31, 165, doi:10.11604/pamj.2018.31.165.15991 (2018).

19 Afakye, K. et al. The Impacts of Animal Health Service Providers on Antimicrobial Use Attitudes and Practices: An Examination of Poultry Layer Farmers in Ghana and Kenya. Antibiotics (Basel, Switzerland) 9, 554, doi:10.3390/antibiotics9090554 (2020).

20 Aywak, D. et al. Pharmacy Practice in Kenya. Can J Hosp Pharm 70, 456–462, doi:10.4212/cjhp.v70i6.1713 (2017).

21 National Council for Law Reporting. (www.kenyalaw.org, 2011).

22 Sakeena, M. H. F., Bennett, A. A. & McLachlan, A. J. Enhancing pharmacists’ role in developing countries to overcome the challenge of antimicrobial resistance: a narrative review. Antimicrobial resistance and infection control 7, 63, doi:10.1186/s13756-018-0351-z (2018).

23 Muloi, D. et al. A cross-sectional survey of practices and knowledge among antibiotic retailers in Nairobi, Kenya. Journal of global health 9, 010412, doi:10.7189/jogh.09.020412 (2019).

24 Fevre, E. M. et al. An integrated study of human and animal infectious disease in the Lake Victoria crescent small-holder crop-livestock production system, Kenya. BMC Infect Dis 17, 457, doi:10.1186/s12879-017-2559-6 (2017).

25 Kenya National Bureaua of Statistics. 2019 Kenya Population & Housing Census (2019).

26 Nowell, L. S., Norris, J. M., White, D. E. & Moules, N. J. Thematic Analysis: Striving to Meet the Trustworthiness Criteria. International Journal of Qualitative Methods 16, 1609406917733847, doi:Artn 1609406917733847 10.1177/1609406917733847 (2017).

27 National Council for Law Reporting. (2012).

28 EMA. European Medicines Agency - Antimicrobial resistance - Recommendations on the use of antibiotics in animals, <http://www.ema.europa.eu/ema/index.jsp?curl=pages/regulation/general/general_content_000639.jsp&mid=WC0b01ac058080a585#chapter2> (2015), Accessed.

29 Horumpende, P. G. et al. Prescription and non-prescription antibiotic dispensing practices in part I and part II pharmacies in Moshi Municipality, Kilimanjaro Region in Tanzania: A simulated clients approach. PloS one 13, e0207465, doi:10.1371/journal.pone.0207465 (2018).

30 de Glanville, W. A. et al. Household socio-economic position and individual infectious disease risk in rural Kenya. Scientific reports 9, 2972, doi:10.1038/s41598-019-39375-z (2019).

31 Tangcharoensathien, V., Chanvatik, S. & Sommanustweechai, A. Complex determinants of inappropriate use of antibiotics. Bull World Health Organ 96, 141–144, doi:10.2471/BLT.17.199687 (2018).

32 Leonard, D. K. The Supply of Veterinary Services - Kenyan Lessons. Agricultural Administration and Extension 26, 219–236, doi:Doi 10.1016/0269-7475(87)90061-4 (1987).

33 Wilkinson, A., Ebata, A. & MacGregor, H. Interventions to Reduce Antibiotic Prescribing in LMICs: A Scoping Review of Evidence from Human and Animal Health Systems. Antibiotics (Basel, Switzerland) 8, doi:10.3390/antibiotics8010002 (2018).

34 WHO. Multicountry public awareness survey. World Health Organization: Geneva, Switzerland, 59 (2015).

35 Caudell, M. A. et al. Towards a bottom-up understanding of antimicrobial use and resistance on the farm: A knowledge, attitudes, and practices survey across livestock systems in five African countries. PloS one 15, e0220274, doi:10.1371/journal.pone.0220274 (2020).

36 Caudell, M. A. et al. Antimicrobial Use and Veterinary Care among Agro-Pastoralists in Northern Tanzania. PloS one 12, e0170328, doi:10.1371/journal.pone.0170328 (2017).

37 Walker, S. Effective antimicrobial resistance communication: the role of information design. Palgrave Communications 5, 24, doi:ARTN 24 10.1057/s41599-019-0231-z (2019).

38 Tadesse, B. T. et al. Antimicrobial resistance in Africa: a systematic review. BMC Infect Dis 17, 616, doi:10.1186/s12879-017-2713-1 (2017).

39 Ampaire, L. et al. A review of antimicrobial resistance in East Africa. African journal of laboratory medicine 5, 432, doi:10.4102/ajlm.v5i1.432 (2016).

40 Lerner, J. S. & Tetlock, P. E. Accounting for the effects of accountability. Psychol Bull 125, 255–275, doi:10.1037/0033-2909.125.2.255 (1999).

41 Kiefe, C. I. et al. Improving quality improvement using achievable benchmarks for physician feedback: a randomized controlled trial. Jama 285, 2871–2879, doi:10.1001/jama.285.22.2871 (2001).

42 Genga, E., Achieng, L., Njiri, F. & Ezzi, M. Knowledge, attitudes, and practice survey about antimicrobial resistance and prescribing among physicians in a hospital setting in Nairobi, Kenya. Knowledge, attitudes, and practice survey about antimicrobial resistance and prescribing among physicians in a hospital setting in Nairobi, Kenya Risk factors for pulmonary tuberculosis treatment failure in rural settings in Benin, West Africa: A cohort study 3 (2017).

43 Chantziaras, I., Boyen, F., Callens, B. & Dewulf, J. Correlation between veterinary antimicrobial use and antimicrobial resistance in food-producing animals: a report on seven countries. Journal of Antimicrobial Chemotherapy 69, 827–834, doi:10.1093/jac/dkt443 (2014).

44 Roth, N. et al. The application of antibiotics in broiler production and the resulting antibiotic resistance in Escherichia coli: A global overview. Poult Sci 98, 1791–1804, doi:10.3382/ps/pey539 (2019).

