## Supplementary material for "A Cross-Sectional Survey of the Knowledge, Attitudes & Practices of Antimicrobial Users and Providers in an Area of High-Density Livestock-Human Population in Western Kenya": Supp Figure 1

### Slide 1
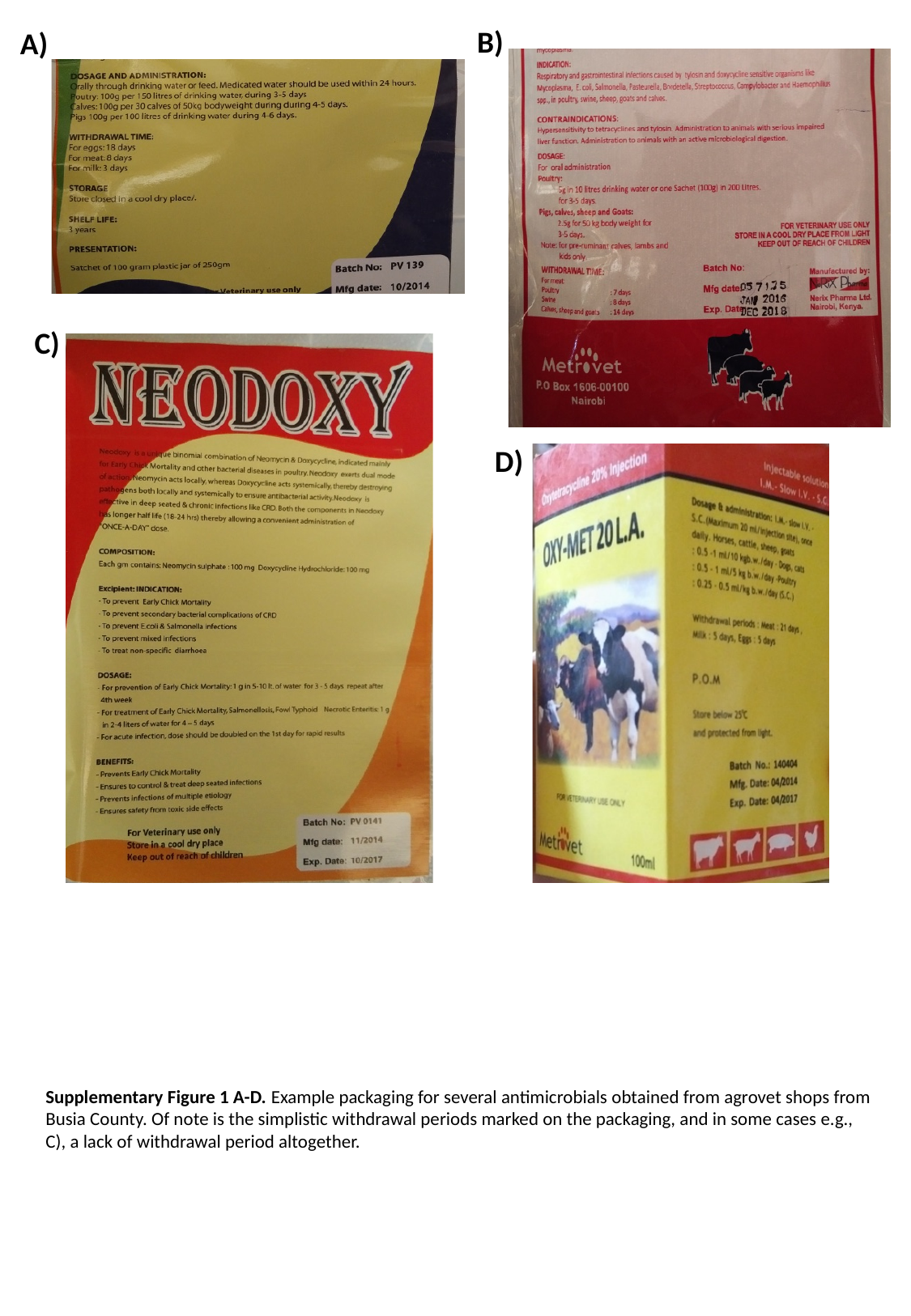

B)
A)
C)
D)
Supplementary Figure 1 A-D. Example packaging for several antimicrobials obtained from agrovet shops from Busia County. Of note is the simplistic withdrawal periods marked on the packaging, and in some cases e.g., C), a lack of withdrawal period altogether.
