## Supplementary material for "A Cross-Sectional Survey of the Knowledge, Attitudes & Practices of Antimicrobial Users and Providers in an Area of High-Density Livestock-Human Population in Western Kenya": Supp Table

**Supplementary Table 1.** Comprehensive list of questions used to interview all farmers, agrovet staff and veterinary professionals.

1. Shop Name/Location

2. Gender

3. Age Group

4. Current Occupation

5. Do you have more than one job?

6. What else do you do?

7. What is the highest degree or level of school you have completed?

8. What sort of establishment is this?

9. What is your job at this outlet?

10. How long have you worked here for?

11. How many other staff work here and what are their roles?

12. How many pharmacies or agrovets are there nearby (within 3km)?

13. Does the Owner work at this outlet?

14. What qualification(s) do they hold?

15. Do you or the owner hold a current license to sell antibiotics for animal use?

16. Do you have a veterinary degree?

17. Have you had specific training in Livestock Health and/or Diseases?

18. Have you ever received training or are you still training to dispense antibiotics for animal use?

19. Please describe the nature of your training.

20. Which of the following products do you sell here?

21. What other products do you sell?

22. Which of the following services do you provide here?

23. What other services do you provide?

24. If you provide diagnostic testing, what specifically do you do?

25. What other diagnostic tests do you perform?

26. Which of the following do you serve most frequently?

27. Which of the following do you serve least frequently?

28. What are the main reasons for customers to come to this shop? To buy/get:

29. What other reasons do people visit the shop for?

30. On average, how many customers do you serve...

31. Do you write prescriptions for antibiotics?

32. How often do you give advice to a customer before they purchase antibiotics?

33. When giving information to customers about withdrawal periods and antibiotic residues, what specific information do you give and where do you get that information from?

34. When recommending/prescribing antibiotics, which of the following pieces of information do you give the customer?

35. What other information do you give to customers?

36. What factors do you take into account when recommending antibiotics to a customer?

37. What other factors do you take into account when recommending antibiotics to a customer?

38. How often does your customer request a specific antibiotic?

39. Have you ever recommended or prescribed antibiotics to a farmer for animals without examining the animals first?

40. Why did you make a recommendation without examining the animals?

41. Once a person has obtained the antibiotics you have recommended or prescribed, how often do you administer them to the animals on their behalf?

42. Once treatment of the animals has begun/been completed, do you perform check-ups on the farm to determine the clinical outcome?

43. Why don't you perform check-ups?

44. Where do you acquire your antibiotics from?

45. Are there any types of antibiotics which you cannot get from your supplier?

46. How do you store your antibiotics?

47. How else do you store your antibiotics?

48. Do you keep a record of all the antibiotics you dispense/prescribe?

49. Please can we see/take a copy?

50. What are the five most commonly prescribed/sold antibiotics?

51. What/which condition was each antibiotic sold to treat?

52. What species was this each antibiotic for?

53. What is the typical dose, frequency and duration of treatment for each antibiotic?

54. What is the price for the customer and what is the price you paid to buy the antibiotics?

55. Are the antibiotics you sell primarily used for:

56. Are there any antibiotics which you are not allowed to recommend/prescribe?

57. Please list the name of each antibiotic and the reason you cannot prescribe them.

58. Are you aware of any specific guidelines which should be consulted when recommending or prescribing antibiotics?

59. Please list any organisations and policies that you are aware of.

60. Are you aware of any critical/important antibiotics which are of high importance to human medicine, but can also be prescribed for animal use?

61. Please give examples of critical/important antibiotics that you are aware of.

62. Have you ever prescribed 3rd or 4th generation Cephalosporins or Fluoroquinolone antibiotics for animal use?

63. Please give details of the clinical scenario in which they were prescribed.

64. If you recommend or prescribe an antibiotic and the farmer returns to you and complains that it hasn't treated the disease, what do you do?

65. Please indicate in the space below what you think/know about what antibiotic resistance is.

66. Have you had any evidence of antibiotic resistance when prescribing or treating animals with antibiotics?

67. What did you do to overcome the antibiotic resistance?

68. What is your job at this shop?

69. How long have you worked here for?

70. How many other staff work here and what are their roles?

71. How many other pharmacies/agrovets are there close by (within 3km)?

72. Does the owner work here?

73. What qualification(s) do they hold?

74. Do you or the owner hold a current license to sell antibiotics for animal use?

75. Have you had specific training in Livestock Health and/or Disease?

76. Have you ever received any training in, or are you still training to dispense antibiotics for animal use?

77. What sort of training have you undertaken or are still undertaking?

78. Do you think you need additional training to help you do your work?

79. What additional training would be of most benefit to your work?

80. What types of product do you sell in this shop?

81. What other types of products do you sell?

82. Do you provide any other services?

83. What other services do you provide?

84. Who are your most frequent customers?

85. Who are your least frequent customers?

86. What are the main reasons for customers to come to this shop? To buy/get:

87. What other reasons do people visit the shop for?

88. On average, how many customers do you serve...

89. Do you fulfil prescriptions for antibiotics here?

90. How often do you give a customer advice before they purchase antibiotics?

91. When giving information to customers about Withdrawal Periods and Residues, what specific information do you give and where do you get that information from?

92. When you sell antibiotics to a customer, which of the following pieces of information do you give them?

93. What other information do you give to customers?

94. What factors do you take into account when recommending antibiotics to a customer?

95. What other factors do you take into account when recommending antibiotics to a customer?

96. How often does your customer make a request a specific antibiotic?

97. If you are unsure of the best antibiotic treatment options for animals, do you refer the customer to a veterinary or other animal healthcare worker?

98. Where do you acquire your antibiotics from?

99. Are there any types of antibiotics which you cannot get from your supplier?

100. How do you store your antibiotics?

101. How else do you store your antibiotics?

102. Do you keep a record of all of the antibiotics you sell?

103. Please can we see/have a copy of this?

104. What are the five most commonly sold antibiotics?

105. What/which condition was each antibiotic sold to treat?

106. What species was each antibiotic for?

107. What is the typical dose, frequency and duration of each treatment?

108. What is the price for the customer and how much did you pay for the antibiotic?

109. Are the antibiotics you sell primarily used for:

110. Are you aware of any specific guidelines which should be consulted when recommending or selling antibiotics?

111. Please list any organisations and policies that you are aware of.

112. Are you aware of any critical/important antibiotics which are of high importance to human medicine, but can also be used for animals?

113. Please give examples of critical/important antibiotics which you are aware of.

114. Do you stock 3rd or 4th generation Cephalosporins or Fluoroquinolones?

115. Have you ever sold 3rd or 4th generation Cephalosporins or Fluoroquinolones?

116. What did you sell and what was the clinical scenario in which they were recommended for?

117. If you recommend and sell an antibiotic to a farmer and the farmer returns and states that his animal has not been cured, what do you do?

118. Please indicate in the space below what you think/know about what antibiotic resistance is.

119. Have you had any evidence of antibiotic resistance when prescribing or treating animals with antibiotics?

120. What did you do to overcome the antibiotic resistance?

121. How long have you worked at this stall for?

122. Are you the owner/does the owner of the stall work here?

123. What qualifications do you/the owner hold?

124. Do you/the owner hold a current license to sell antibiotics for animal use?

125. Have you had specific training in Livestock Health and/or Disease?

126. Have you ever received any training in, or are you still training to sell antibiotics for animal use?

127. What sort of training have you undertaken or are still undertaking?

128. Do you think you need additional training to help you do your work?

129. What additional training would be of most benefit to your work?

130. What types of product do you sell at this stall?

131. What other types of product do you sell here?

132. Do you provide any services to customers?

133. What other services do you provide?

134. Who are your most frequent customers?

135. Who are your least frequent customers?

136. What are the main reasons for customers to come to this stall? To buy/get:

137. What other reasons do people visit this shop for?

138. On average, how many customers do you serve...

139. How often do you give a customer advice before they purchase antibiotics?

140. When giving information to customers about Withdrawal Periods and Residues, what specific information do you give and where do you get that information from?

141. When you sell antibiotics to a customer, which of the following pieces of information do you give them?

142. What other information do you give to customers?

143. What factors do you take into account when recommending antibiotics to a customer?

144. What other factors do you take into account when recommending antibiotics to a customer?

145. How often does your customer make a request for a specific antibiotic?

146. If you are unsure of the best antibiotic treatment options for animals, do you refer the customer to a veterinary or other animal healthcare worker?

147. Where do you acquire your antibiotics from?

148. Are there any types of antibiotics which you cannot get from your supplier?

149. How do you store your antibiotics?

150. How else do you store your antibiotics?

151. Do you keep a record of all of the antibiotics you sell?

152. Please can we see/have a copy of this?

153. Image

154. What are the five most commonly sold antibiotics?

155. What/which condition was each antibiotic sold to treat?

156. What species was each antibiotic for?

157. What is the typical dose, frequency and duration of each treatment?

158. What is the price for the customer and how much did you pay for the antibiotic?

159. Are the antibiotics you sell primarily used for:

160. Are you aware of any specific guidelines which should be consulted when recommending or selling antibiotics?

161. Please list any organisations and policies that you are aware of.

162. Are you aware of any critical/important antibiotics which are of high importance to human medicine, but can also be used for animals?

163. Please give examples of critical/important antibiotics which you are aware of.

164. Do you stock 3rd or 4th generation Cephalosporins or Fluoroquinolones?

165. Do you ever sold 3rd or 4th generation Cephalosporins or Fluoroquinolones?

166. What did you sell and what was the clinical scenario in which they were recommended for?

167. If you sell an antibiotic to a farmer and the farmer returns and states that his animal has not been cured, what do you do?

168. Please indicate in the space below what you think/know about what antibiotic resistance is.

169. Have you had any evidence of antibiotic resistance when selling antibiotics for animals?

170. What did you do to overcome the antibiotic resistance?

171. Which types of animal do you keep on your farm?

172. What other animals do you keep here?

173. How many of each animal do you have?

174. What is the main purpose of your animals?

175. How many of your animals are used for breeding?

176. How many of your animals are used for animal products for sale (milk/meat/eggs etc.)

177. How many of your animals are used for animal products for your own consumption (milk/meat/eggs etc.)

178. Do you treat your animals with antibiotics?

179. Can you estimate how often you treat your animals with antibiotics e.g. weekly, monthly, yearly?

180. Which of the following reasons do you treat your animals with antibiotics for?

181. Have any of your animals required antibiotic treatment in the last year?

182. Please list the illnesses and which types of antibiotics, you used to treat your animals with.

183. Do you ever purchase commercial feeds containing...

184. When purchasing commercial feeds containing antibiotics, what is it mainly used for?

185. Do you keep a record of all of the antibiotics you treat your animals with?

186. Can we have a copy?

187. Do you seek the advice of a vet before purchasing antibiotics?

188. If you do not seek advice from a vet before purchasing antibiotics, do you seek advice elsewhere?

189. Which other sources do you consult?

190. How often do your request a specific antibiotic from an agrovet or pharmacy?

191. Why do you request specific antibiotics?

192. How do you know to request that specific antibiotic?

193. Where do you purchase your antibiotics from?

194. What is the reason for choosing this place?

195. How many pharmacies or agrovets are there nearby (within 3km)?

196. What are the main factors when purchasing antibiotics?

197. What other factors do you take into consideration when purchasing antibiotics?

198. Do you have an issue with the amount of antibiotics you need to purchase e.g. you have two sheep, but you can only buy packs to treat ten sheep?

199. Do you use any extra antibiotics for any other animal species or do you store them?

200. How do you store any extra antibiotics which do you do not use?

201. How else do you store any antibiotics which you do not use?

202. Once you have purchased the antibiotic(s), do you follow the recommended dosage instructions?

203. If you treat an animal with antibiotics and it appears to get better before the end of the full treatment, do you stop giving it antibiotics and save them for later?

204. Can you estimate how often you treat your animals e.g. daily, weekly, monthly or yearly?

205. How do you work out the correct dosage for each animal you need to treat with antibiotics?

206. What other ways do you use to estimate the correct dose of antibiotics?

207. How do you give the antibiotics to the pigs?

208. How do you give the antibiotics to the chickens?

209. How do you give the antibiotics to the cattle?

210. How do you give the antibiotics to the sheep?

211. How do you give the antibiotics to the goats?

212. If you have selected 'other' for any of the questions asking how you give the antibiotics to your animals, please write below the name of the animals and how else you give antibiotics to them.

213. Do you always give the recommended dosage to your animals?

214. Why don't you always give the recommended dosage to your animals??

215. Imagine the following clinical scenario: You have a 10 cattle, but you need antibiotics for just 1 of them. You check the dose instructions on the antibiotics packaging but it does not allow for just 1 animal to be treated. What do you do?

216. If you purchase one type of antibiotic to treat the animals with and it does not work, what do you do?

217. What do you know about antibiotic resistance?

218. Have you had any evidence of antibiotic resistance when giving antibiotics to your animals?

219. What do you think causes antibiotic resistance?

220. What did you do to overcome the antibiotic resistance?

221. What do you know about antibiotic withdrawal periods?

222. Do you sell any animal products (meat/cheese/milk/eggs) whilst the animals are on antibiotics or have recently finished treatment?

223. Do you sell consume any animal products (meat/cheese/milk/eggs) whilst the animals are on antibiotics or have recently finished treatment?

224. If you sell milk obtained from your animals, do you pasteurise or boil your milk before selling it?

225. If you consume milk obtained from your animals, do you pasteurise or boil your milk before consuming it?

226. Have you ever purchased antibiotics intended for human consumption from a pharmacy or doctor to give to your animals?

227. Please give details of what the antibiotic was and why it was used instead of a veterinary antibiotic.
